# Default-filled outcome labels in a deployed cognitive-screening programme: an operator-level audit and the construction of twenty-four language-model arms

**DOI:** 10.64898/2026.08.28.26361585

**Authors:** Jun Ji, Zhigang Sun, Xiaofang Ying, Jinyu Hao, Zhiqiang Fu, Dongmei Shi, Xiaoming Kong, Yijie Xu, Xiaojuan Zhang, Xiaoli Du, Zhiyan Zhang, Xinhong Liu, Ping Lin, Huali Wang

## Abstract

**Background:** Routine service databases are attractive sources of training labels for clinical prediction models, but the processes that write those labels are rarely audited before the labels are used. In a deployed community cognitive-screening programme we audited the routine cognitive-status label, built a matrix of twenty-four model arms over the same patients under a specialist reference standard, and measured what each supervision choice bought or cost.

**Methods:** The study cohort is the 672 individuals whose cognitive status was recorded by a titled (attending-or-above) physician, that record being the reference standard; after holding out one institution entirely, a development panel of 642 individuals at 38 institutions. The routine cognitive-status label these individuals also carry was first audited at the operator level: for each data-entry account we counted diagnoses entered and the proportion recording any impairment, and we tested a competing bulk-timestamp explanation. Twenty-four arms span the supervision choices such a programme faces: an incumbent 21-variable logistic regression; local language models (Qwen2.5-1.5B/3B, Qwen3-4B/8B) zero-shot, with chain-of-thought, fine-tuned on physician labels, fine-tuned on routine labels with and without decontamination, or fine-tuned on a proxy scale-band task; preference-optimised (DPO) and reinforcement-trained (GRPO) variants; a proprietary frontier model queried zero-shot; and knowledge distillation of that frontier model into the regression and into the local 4B, using 943 teacher-labelled records drawn from the programme’s unlabelled pool. All arms are scored out-of-fold under one five-fold split grouped on registry-resolved institution clusters (no cluster spans a fold); paired contrasts use a 2,000-draw cluster bootstrap.

**Results:** 181 operator accounts — each entering at least 100 diagnoses with zero recorded impairments — account for 45,315 rows, or 40.5% of the outcome column; recorded impairment falls monotonically with account volume (15.7% for 1–9 rows down to 0.7% for 500–999); a bulk-timestamp explanation was tested and refuted, identifying the write-time column as a migration artefact. Under the specialist standard, no locally fine-tuned arm beat the incumbent regression (AUROC 0.926): physician-label SFT reached 0.924 (4B), DPO 0.881 and GRPO 0.789, and the pre-registered two-stage proxy-then-RL recipe was *worse* than its single-stage contaminated baseline (−0.030, 95% CI −0.077 to −0.004). Chain-of-thought reduced discrimination at every size (−0.072, −0.080, −0.041 at 1.5B/3B/4B; −0.012, n.s., at 8B). The frontier model scored 0.932 (vs. regression +0.007, n.s.). The distilled 4B reached 0.940 — above the incumbent (+0.014, 0.004 to 0.031) and above its own teacher (+0.008, 0.001 to 0.017) — with near-teacher calibration; it reached the teacher’s level by 50 teacher labels and changed little beyond 200.

**Conclusions:** The audit and the arm matrix support one deployment recipe: audit the routine label at the operator level before training on it; do not expect fine-tuning, preference optimisation, or reinforcement learning on a few hundred specialist cases to beat a well-calibrated regression; and if a frontier model is available but undeployable, spend a bounded number of queries on it as a labelling instrument and distil. A companion paper uses these frozen predictions to quantify how evaluation design choices compare with model choice.

## 1 Introduction

Deployed screening programmes accumulate two very different assets. The first is large and cheap: a routine database in which every service contact writes a structured outcome — here, 112,023 cognitive-status rows across a community cognitive-screening programme. The second is small and expensive: the subset of individuals whose status was recorded by a specialist — here, 672 individuals diagnosed by titled (attending-or-above) physicians. The default machine-learning move is to train on the first and hope it approximates the second. This paper reports, for one deployed programme, why that default fails and what the alternatives actually deliver.

The failure is not random noise. Auditing the routine outcome column at the *operator* level — that is, by who typed each row — shows that 181 accounts, each entering at least one hundred diagnoses without ever recording an impairment, wrote 40.5% of the column. The pattern survives a deliberately sought competing explanation, bulk timestamp auto-fill, which the data refute. Because the contamination is concentrated in identifiable accounts rather than spread at random, it cannot be averaged away by sample size; but it *can* be detected from provenance metadata that most service databases already store.

Against this backdrop we constructed twenty-four model arms over the same patients and the same specialist reference standard: the programme’s incumbent 21-variable logistic regression, local open-weight language models at four sizes under seven supervision regimes, a proprietary frontier model, and knowledge distillation of the frontier model into both the regression and the strongest local model. The matrix includes a pre-registered two-stage protocol (proxy pre-training, then reinforcement alignment on specialist labels) and two deliberate controls: a chain-of-thought ladder and a known label-space-mismatch proxy arm.

The contributions are:

1. An operator-level contamination audit with a tested-and-refuted competing mechanism, reproducible from provenance metadata alone.
2. A training report for twenty-four arms under a single leakage-free evaluation design — folds grouped on registry-resolved institution clusters, a cluster bootstrap for every interval — with the pre-registered claims adjudicated.
3. A costed distillation recipe whose entire teacher budget is under one thousand queries, reported together with the label-budget curve, a teacher-noise measurement, and a train–test overlap sensitivity analysis.

A companion paper [22] freezes the predictions made here and audits the *evaluation* side: how much the outcome definition, the clustering structure, and the metric change conclusions relative to model choice. The two papers share one set of frozen artefacts; every number in both is regenerated by released scripts.

## 2 Related work

Label quality in routine health data is a known hazard: reuse of electronic health record data inherits the care processes that produced it [1,3,4], and learning under label noise is well studied when the noise is random or class-conditional [2]. The contamination documented here is neither. It is concentrated in identifiable operator accounts, which is precisely what makes it both detectable and resistant to averaging.

Large language models have been proposed as annotators in place of human raters [9,10]. We use a frontier model in that role — as a labelling instrument rather than as a predictor — and distil its verdicts into deployable students [11,12].

On the training side we use parameter-efficient fine-tuning [13], direct preference optimisation [14], and group-relative policy optimisation [15]. On the evaluation side we use calibration diagnostics [16,17], decision curves [18], and cluster-aware inference for multi-centre data [19], reported under TRIPOD+AI [20].

### 3 Data, labels, and the contamination audit

#### 3.1 Setting

Data arise from a community cognitive-screening programme in routine use across mainland China: residents complete the AD-8 informant interview [5] and a CSI-D-derived cognitive test [6,7] — the Hong Kong Brief Cognitive Test (HKBC) [8] — and a deployed 21-variable logistic regression flags residents for referral. Records are pseudonymised at source; the data custodian is the Beijing Medical Award Foundation. Individuals gave informed consent electronically through the programme’s information system at the point of service. The study — a retrospective statistical analysis of de-identified records with no clinical intervention — was approved by the Ethics Committee of the Medical College of Qingdao University (approval no. QDU-HEC-2025431).

#### 3.2 Three labels

**The routine label** is the latest structured cognitive diagnosis in the programme database: 112,023 rows entered by 2,597 operator accounts. Its class decomposition is extreme. Of 105,893 assessed individuals, 103,688 are recorded normal, 2,197 unspecified cognitive impairment, 1 mild cognitive impairment, and 7 dementia. We therefore write “cognitive impairment as recorded” throughout, and never treat this column as a clinical construct.

**The specialist (reference) label** is the recorded diagnosis of a titled (attending-or-above) physician. Assembling every such individual, then excluding records missing the instruments, test-team accounts, and one placeholder row, yields 672 individuals. The registry contains exactly four distinct diagnosis strings — “cognitively normal”, “cognitive impairment”, and two specific dementia diagnoses — recorded by attending (419), associate-chief (156) and chief (67) physicians on the development panel, all before any model arm in this study existed. The cluster identifier is the institution, resolved per individual from the platform’s team registry, which is definitive where the physician-typed name is not. One institution (Peking University Sixth Hospital) was held out entirely for a held-out-site check, leaving a **development panel of 642 individuals at 38 institutions** (259 impaired, 40.3%). The panel is case-mix selected relative to the programme (40.3% vs. 2.6% recorded impairment), so pooled discrimination here is not a population estimate — a point the companion paper quantifies.

**The proxy label** is the HKBC scale band (mild vs. severe impairment range), available for roughly 43,000 programme records. It is abundant, but by design occupies a different label space from the clinical verdict; the arm trained only on it serves as a known label-space-mismatch control.

#### 3.3 Operator-level audit: method

For each of the 2,597 operator accounts we counted (i) diagnoses entered and (ii) the proportion recording any impairment, then stratified accounts by volume. The detection rule flags accounts with at least 100 diagnoses and *zero* recorded impairments: under the programme’s observed base rate, hundreds of consecutive negatives from one account is implausible as case mix.

To separate default entry from genuine screening of low-risk populations we tested a competing explanation. If defaults were written by bulk processes, they should cluster in write-timestamps. The densest single second in the table contains 26,694 rows and spans *all* operator classes — titled physicians’ rows are *more* timestamp-concentrated than untitled accounts’ rows. This identifies the write-time column as a one-off migration artefact rather than an auto-fill signature, and disqualifies that column, rather than the operator dimension, from carrying any inference. Consequently no temporal claim is made anywhere in this study.

## 4 The twenty-four arms

### 4.1 Shared representation and readout

Every language-model arm sees the identical rendered input: the 8 AD-8 informant items, 9 CSI-D cognitive items, immediate recall, age, sex, and education, rendered item-by-item by one shared renderer — character-identical across arms and stages — with the instruction to answer “cognitively normal” or “cognitively impaired” (in Chinese). The predicted probability is the restricted softmax over the two answer tokens. The incumbent regression uses the same 21 variables. Local models are Qwen2.5-1.5B-Instruct, Qwen2.5-3B-Instruct, Qwen3-4B-Instruct-2507 and Qwen3-8B [23,24], loaded in 4-bit NF4; all fine-tuning is LoRA-based on a single consumer GPU with 8 GB of memory. The frontier model (gpt-5.6-pro) is queried through a gateway that exposes neither token probabilities nor sampling controls, so its probability is elicited as a verbalised integer 0–100 and divided by 100. The referral threshold deployed by the programme is 0.10; the companion paper additionally analyses re-derived thresholds.

### 4.2 The arm matrix

Table 2 lists all twenty-four arms as (supervision source × objective × start × size). The design logic is as follows.

- **a5_lr — incumbent architecture**. The 21-variable logistic regression, re-fitted cross-fitted to physician labels. It characterises the architecture, not the frozen production instance, and it is the bar every learned arm must clear, because it uses exactly the information the language models see in prompt.
- **a0 / a0c — routine label, single stage (1.5B)**. SFT on the rendered routine-label corpus (5,752 training rows). a0c first drops rows entered by high-volume zero-positive accounts (at least 200 diagnoses with zero impairments: 54 accounts, 27,616 database rows), leaving 4,616 training rows (decontamination).
- **a1 — proxy task only (1.5B)**. The Stage-1 adapter alone: SFT on the HKBC scale-band task (30,081 training rows after excluding every reference-cohort participant from the ~43,000 rendered records), never shown a clinical verdict — the known label-space-mismatch control.
- **a2 family — the pre-registered two-stage recipe (1.5B)**. Stage-1 proxy adapter, then reinforcement alignment (GRPO) on specialist labels with a cost-weighted reward (λ penalises missed cases); λ ∈ {3, 9, 27} (a2_lam3, a2, a2_lam27).
- **a3 family — specialist labels from scratch (1.5B, 4B)**. SFT from a randomly initialised LoRA on the physician labels. At 4B the same start, folds, and hyperparameters also run under DPO (a3dpo) and GRPO (a3grpo), giving a clean three-way supervised-vs-preference-vs-reinforcement comparison. DPO here has no human preference data: with two actions and preference fixed by the label, it reduces to a logistic loss on the reference-anchored log-odds margin.
- **a4 — proxy then specialist SFT (1.5B)**. The critical ablation for the two-stage claim: it shares a2’s start and hyperparameters, replacing the RL objective with cross-entropy. It inherits the RL learning rate (2 × 10^−5^) by design; a4_lr2e4 re-runs it at the SFT-conventional 2 × 10^−4^ as sensitivity.
- **a6z / a6c — zero-shot ladder (1.5B, 3B, 4B, 8B)**. The no-training baselines, without (a6z) and with (a6c) an explicit chain-of-thought pass. a6c is a diagnostic control, not a deployment candidate.
- **a7 — frontier zero-shot**. gpt-5.6-pro on every panel individual, same rendered input.
- **a5* — distilled regression**. The incumbent architecture re-fitted, per fold, to the frontier model’s thresholded calls on that fold’s training patients; physician labels are used only for evaluation.
- **a8distil — distilled 4B**. The twenty-fourth arm, trained not on panel patients at all but on teacher-labelled records from the programme’s unlabelled pool (Section 4.3).

### 4.3 Distillation from the frontier teacher

The unlabelled pool is the programme’s routine records with complete instrument responses, excluding every reference-cohort participant: 43,553 rows, minus 510 sharing a participant with the reference cohort, minus 220 incomplete, leaving 42,823 rows that collapse to 15,127 distinct rendered patterns. Patterns were sampled under a label-free risk stratification (AD-8 endorsements plus CSI-D errors, capped at 8, nine bands) with stored inclusion probabilities, under a nested design whose prefixes are themselves valid designs; the plan budgeted 5,000 teacher calls covering 64.5% of pool rows directly. The deployed arm’s training set is the design’s first 944 labelled patterns — well inside the plan — of which 943 were used (one was dropped because its subject later entered the reference standard). Labelling later continued to 1,944 patterns under the same design for other analyses, but every result reported here uses the 943. Throughput on the first 472-call tranche was a median 12.1 s per call with zero unparseable responses. Teacher noise was quantified by repeat-labelling 100 patterns: 24% of repeats identical, 72% within ±0.05, mean absolute range 0.053 (maximum 0.25) — the noise a student averages over.

The student is the 4B with the physician-label arm’s initialisation and hyperparameters (LoRA, lr 2 ×10^−5^, 2 epochs, batch 2, seed 42), trained with weighted cross-entropy over the two answer tokens against the teacher’s verbalised scores; the weights are normalised inverse-inclusion-probability (Horvitz–Thompson) weights (mean 1, maximum 44.8). The two 4B SFT arms differ only in supervision source, records, and weighting. Training takes about seven minutes on the 8 GB GPU. Because the pool is participant-disjoint from the panel, one distilled model scores every panel individual out-of-sample, and no physician label entered training. Rendered inputs are discrete patterns, so identical inputs across participants remain possible: 16 of the 944 teacher patterns (1.7%) match a panel individual’s rendered input, covering 18 of 642 panel individuals. Excluding them moves the distilled arm’s pooled AUROC from 0.940 to 0.940 and its fixed-threshold ΔNB100 from +3.13 to +3.15, so no reported result rests on the overlap.

### 4.4 Folds, evaluation, and statistics

All fold-trained arms use one five-fold split of the 672, stratified by outcome and grouped on the registry-resolved institution clusters, so that no cluster spans a fold boundary — an assertion checked programmatically against the final cluster identities. (An earlier lineage of this project compared raw name strings and leaked; the released folds are re-cut, and the leak check reports zero affected individuals.) Every arm scores every development-panel individual out-of-fold; fold-independent arms — zero-shot, frontier, distilled — are simply scored once.

Discrimination is AUROC. Decision value is net benefit at the deployed threshold 0.10, reported as ΔNB100, the difference per 100 individuals against the best trivial policy. Calibration is summarised by expected calibration error, calibration slope, and the fraction of outputs below 0.01 or above 0.99 (saturation). Paired contrasts are computed within bootstrap replicate under a 2,000-draw cluster bootstrap resampling institutions (seed 20260803).

The realised design is heavily clustered: empirical design effect median 4.06 across arms (range 2.24– 8.24), effective sample size near 158 of 642, outcome intraclass correlation 0.360. All contrasts are exploratory; where multiplicity is corrected, the companion paper’s Benjamini–Hochberg families apply. Evaluation of the held-out institution (58 individuals with complete instrument responses of 241 diagnosed; reference label from any operator, 52 of 58 cases) uses an individual-level bootstrap and is an exploratory same-programme check, not external validation.

## 5 Results

### 5.1 Forty percent of the outcome column is default-filled

Recorded impairment falls monotonically with operator volume (Table 1): accounts entering 1–9 diagnoses record impairment at 15.7%; 10–49 at 3.4%; 100–499 at 2.1%; 500–999 at 0.7%. The 181 accounts with at least 100 diagnoses and zero impairments wrote 45,315 rows, or 40.5% of the column; six accounts each entered over 1,000 diagnoses without one impairment (largest: 3,960). Excluding the flagged accounts raises observed programme impairment from 2.60% to 4.37%. The bulk-timestamp explanation is refuted (Section 3.3): the contamination is an operator-behaviour signature, detectable from provenance metadata alone, and the write-time column is a migration artefact unusable for temporal inference.

**Table 1.** Recorded impairment by data-entry account volume (112,023 diagnosis rows, 2,597 operator accounts).

| Diagnoses entered per account | Accounts | Rows | Recorded impairment |
| --- | --- | --- | --- |
| 1–9 | 1,368 | 3,106 | 15.7% |
| 10–49 | 723 | 18,870 | 3.4% |
| 50–99 | 227 | 16,240 | 3.5% |
| 100–499 | 248 | 44,227 | 2.1% |
| 500–999 | 22 | 14,144 | 0.7% |
| ≥1,000 | 9 | 15,726 | 1.2% |

*Notes.* Flagged accounts (≥100 diagnoses, zero impairments): 181 accounts, 45,315 rows (40.5% of the column); six of them entered over 1,000 diagnoses each without one recorded impairment. Excluding flagged accounts moves observed programme impairment from 2.60% to 4.37%. The bulk-timestamp competing explanation is refuted (Section 3.3).

**Table 2.** The twenty-four arms: supervision source, objective, start, and size.

| Code | Arm (as named in the companion paper) | Base model | Start | Objective | Supervision source |
| --- | --- | --- | --- | --- | --- |
| a5_lr | Incumbent logistic regression | — (21-variable LR) | — | logistic | physician labels (cross-fitted) |
| a5* | Regression refitted to frontier calls | — (21-variable LR) | — | logistic | teacher calls on training folds |
| a7 | Frontier model, zero-shot | gpt-5.6-pro | — | — | none |
| a8distil | Local 4B, distilled from frontier teacher | Qwen3-4B | random LoRA | weighted CE | 943 teacher-labelled pool records |
| a3 (1.5B/4B) | Local, physician labels | Qwen2.5-1.5B / Qwen3-4B | random LoRA | CE | physician labels |
| a3dpo | Local 4B, preference-optimised | Qwen3-4B | random LoRA | DPO | physician labels |
| a3grpo | Local 4B, RL (GRPO) | Qwen3-4B | random LoRA | GRPO | physician labels |
| a4 | Local 1.5B, physician labels (proxy start) | Qwen2.5-1.5B | Stage-1 proxy | CE | physician labels |
| a4_lr2e4 | — same, higher learning rate | Qwen2.5-1.5B | Stage-1 proxy | CE | physician labels |
| a2 ($\lambda=9$) | Local 1.5B, reinforcement-trained | Qwen2.5-1.5B | Stage-1 proxy | GRPO | physician labels |
| a2_lam3 / a2_lam27 | — same, $\lambda=3$ / $\lambda=27$ | Qwen2.5-1.5B | Stage-1 proxy | GRPO | physician labels |
| a0 | Local 1.5B, routine labels | Qwen2.5-1.5B | random LoRA | CE | routine labels (5,752 rows) |
| a0c | — same, decontaminated | Qwen2.5-1.5B | random LoRA | CE | routine labels excl. zero-positive accounts (4,616 rows) |
| a1 | Local 1.5B, proxy scale-band task (control) | Qwen2.5-1.5B | random LoRA | CE | HKBC scale bands (30,081 rows) |
| a6z (1.5B/3B/4B/8B) | Zero-shot | Qwen2.5-1.5B/3B, Qwen3-4B/8B | — | — | none |
| a6c (1.5B/3B/4B/8B) | Chain-of-thought (control) | Qwen2.5-1.5B/3B, Qwen3-4B/8B | — | — | none |

*Notes.* All local training is LoRA-based, 4-bit NF4, on a single 8 GB consumer GPU. a2 vs. a4 isolates the objective (same start, folds, lr, epochs, batch, seed); a3 vs. a3dpo vs. a3grpo isolates the objective at 4B from a shared random start; a0 vs. a0c isolates decontamination; a1 is a deliberate label-space-mismatch control. The a4 family inherits the RL learning rate $2 \times 10^{-5}$ by design; a4\_lr2e4 is the sensitivity run at $2 \times 10^{-4}$ . CE = cross-entropy.

### 5.2 Arm-by-arm results

Table 3 reports every arm. Four headline patterns emerge.

**Table 3.** All arms on the development panel (*n* = 642, 38 institutions; 2,000-draw cluster bootstrap; fixed referral threshold 0.10). Ordered by pooled AUROC.

| Arm | Pooled AUROC (95% CI) | ECE | Sat. | Sens | Spec | Refer. | $\Delta\text{NB100}$ |
| --- | --- | --- | --- | --- | --- | --- | --- |
| Local 4B, distilled from frontier teacher | 0.940 (0.898–0.963) | 0.045 | 16% | 0.965 | 0.684 | 58% | +3.13 |
| Regression refitted to frontier calls | 0.934 (0.890–0.959) | 0.093 | 74% | 0.900 | 0.807 | 48% | +1.30 |
| Frontier model, zero-shot | 0.932 (0.886–0.960) | 0.041 | 15% | 0.961 | 0.679 | 58% | +2.94 |
| Local 4B, zero-shot | 0.929 (0.889–0.955) | 0.309 | 64% | 0.981 | 0.446 | 73% | +2.18 |
| Incumbent logistic regression | 0.926 (0.873–0.956) | 0.039 | 12% | 0.958 | 0.629 | 61% | +2.46 |
| Local 1.5B, physician labels (proxy start) | 0.926 (0.878–0.953) | 0.081 | 51% | 0.931 | 0.702 | 55% | +1.85 |
| Local 8B, zero-shot | 0.925 (0.886–0.949) | 0.391 | 31% | 1.000 | 0.000 | 100% | 0.00 |
| Local 4B, physician labels | 0.924 (0.870–0.955) | 0.102 | 68% | 0.915 | 0.770 | 51% | +1.68 |
| Local 1.5B, physician labels | 0.924 (0.878–0.952) | 0.085 | 58% | 0.938 | 0.692 | 56% | +2.09 |
| Local 8B, chain-of-thought | 0.913 (0.857–0.943) | 0.344 | 63% | 0.996 | 0.065 | 96% | +0.28 |
| Local 1.5B, decontaminated routine labels | 0.907 (0.844–0.943) | 0.536 | 3% | 1.000 | 0.000 | 100% | 0.00 |
| Local 1.5B, routine labels | 0.899 (0.822–0.939) | 0.278 | 0% | 1.000 | 0.000 | 100% | 0.00 |
| Local 1.5B, zero-shot | 0.890 (0.827–0.929) | 0.555 | 7% | 1.000 | 0.000 | 100% | 0.00 |
| Local 4B, chain-of-thought | 0.888 (0.827–0.923) | 0.137 | 80% | 0.911 | 0.786 | 50% | +1.63 |
| Local 3B, zero-shot | 0.886 (0.834–0.934) | 0.478 | 6% | 1.000 | 0.000 | 100% | 0.00 |
| Local 4B, preference-optimised (DPO) | 0.881 (0.788–0.931) | 0.131 | 99% | 0.880 | 0.817 | 46% | +0.59 |
| Local 1.5B, physician labels (higher LR) | 0.878 (0.794–0.929) | 0.160 | 33% | 0.919 | 0.616 | 60% | +0.81 |
| Local 1.5B, reinforcement-trained ($\lambda = 9$, default) | 0.869 (0.774–0.920) | 0.157 | 97% | 0.849 | 0.796 | 46% | −0.80 |
| Local 1.5B, reinforcement-trained ($\lambda = 3$) | 0.865 (0.786–0.918) | 0.158 | 95% | 0.819 | 0.783 | 46% | −2.13 |
| Local 1.5B, reinforcement-trained ($\lambda = 27$) | 0.841 (0.755–0.900) | 0.249 | 98% | 0.938 | 0.606 | 61% | +1.52 |
| Local 1.5B, chain-of-thought | 0.818 (0.722–0.863) | 0.303 | 13% | 0.992 | 0.180 | 89% | +0.88 |
| Local 3B, chain-of-thought | 0.806 (0.712–0.856) | 0.229 | 55% | 0.946 | 0.567 | 64% | +1.57 |
| Local 1.5B, proxy scale-band task (control) | 0.792 (0.663–0.862) | 0.482 | 1% | 1.000 | 0.000 | 100% | 0.00 |
| Local 4B, RL with reasoning (GRPO) | 0.789 (0.708–0.890) | 0.248 | 89% | 0.938 | 0.606 | 61% | +1.52 |

*Notes.* ECE = expected calibration error. Sat. = saturation, the fraction of outputs below 0.01 or above 0.99 (the companion paper’s calibration figure uses 0.02/0.98 bounds, hence its slightly higher percentages). Sens and Spec are at the fixed threshold. Refer. = fraction flagged at threshold 0.10; 100% referral means every output exceeds the threshold and the arm recovers the trivial refer-everyone policy ( $\Delta\text{NB100} = 0$ against the best trivial rule). $\Delta\text{NB100}$ = net benefit per 100 individuals against the best trivial policy at the deployed threshold ( $\lambda = 9$ ). Within-institution AUROC, tuned-threshold decision value, and all multiplicity-corrected contrasts are reported in the companion paper [22] from the same frozen predictions.

**Table 4.** Key paired contrasts (pooled AUROC; 2,000-draw cluster bootstrap; paired within replicate).

| Question | Contrast | $\Delta$ AUROC (95% CI) | Verdict |
| --- | --- | --- | --- |
| Does two-stage beat single-stage on contaminated labels? (C1) | $a2 - a0$ | $-0.030$ ($-0.077$ to $-0.004$) | refuted (opposite) |
| Does RL beat continued SFT from the same start? (C2) | $a2 - a4$ | $-0.057$ ($-0.122$ to $-0.022$) | refuted |
| — at the SFT-conventional learning rate (C2') | $a2 - a4\_lr2e4$ | $-0.009$ ($-0.067$ to $+0.027$) | indeterminate |
| Does proxy pre-training beat specialist-only? (C3) | $a2 - a3$ (1.5B) | $-0.055$ ($-0.121$ to $-0.018$) | refuted |
| Does decontamination recover performance? (C5) | $a0c - a0$ | $+0.008$ ($-0.005$ to $+0.026$) | indeterminate |
| Did 30k routine labels beat no training? | $a0 - a6z$ (1.5B) | $+0.009$ ($-0.014$ to $+0.024$) | no gain |
| Wrong-label-space fine-tuning vs. no training | $a1 - a6z$ (1.5B) | $-0.098$ ($-0.180$ to $-0.049$) | worse than nothing |
| DPO vs. SFT, same start (4B) | $a3dpo - a3$ | $-0.043$ ($-0.095$ to $-0.014$) | DPO worse |
| GRPO vs. SFT, same start (4B) | $a3grpo - a3$ | $-0.135$ ($-0.199$ to $-0.043$) | GRPO worse |
| Chain-of-thought net effect, 1.5B | $a6c - a6z$ | $-0.072$ ($-0.116$ to $-0.048$) | penalty |
| — 3B | $a6c - a6z$ | $-0.080$ ($-0.162$ to $-0.037$) | penalty |
| — 4B | $a6c - a6z$ | $-0.041$ ($-0.085$ to $-0.008$) | penalty |
| — 8B | $a6c - a6z$ | $-0.012$ ($-0.043$ to $+0.009$) | indistinguishable |
| Any fine-tuned local vs. incumbent regression | $a3$ (4B) $- a5\_lr$ | $-0.002$ ($-0.018$ to $+0.022$) | tie (best case) |
| Frontier vs. incumbent regression | $a7 - a5\_lr$ | $+0.007$ ($-0.005$ to $+0.026$) | not separable |
| Distilled 4B vs. incumbent regression | $a8distil - a5\_lr$ | $+0.014$ ($+0.004$ to $+0.031$) | distilled better |
| Distilled 4B vs. its own teacher | $a8distil - a7$ | $+0.008$ ($+0.001$ to $+0.017$) | student above teacher |
| Distilled 4B vs. physician-label 4B | $a8distil - a3$ (4B) | $+0.016$ ($+0.0004$ to $+0.032$) | teacher labels ≥ physician labels |

*Notes.* Contrast signs are reported in the stated direction; intervals are percentile cluster-bootstrap intervals. All contrasts are exploratory; the companion paper reports Benjamini–Hochberg $q$ -values within its contrast families. C-numbers refer to the pre-registered protocol claims (Section 4.2).

#### No local training beat the incumbent regression

The regression’s pooled AUROC is 0.926 (95% CI 0.873–0.956). Physician-label SFT ties it at every size that can be estimated — 4B 0.924 (Δ vs. a5_lr −0.002, 95% CI −0.018 to +0.022), 1.5B 0.924, proxy-then-SFT 0.926 — and nothing else comes close. The two-stage recipe and all its ablations sit strictly below.

#### The pre-registered claims failed — one of them in the opposite direction

Under leakage-free folds, the two-stage proxy-then-RL arm (a2) is *worse* than the single-stage arm trained on contaminated routine labels (C1: a2 − a0 = −0.030, −0.077 to −0.004); worse than continued supervised fine-tuning from the same start (C2: a2 − a4 = −0.057, −0.122 to −0.022); and worse than specialist-only training (C3: a2 − a3 = −0.055, −0.121 to −0.018). Decontamination recovered a small, inconclusive amount (C5: a0c − a0 = +0.008, −0.005 to +0.026). The λ sweep did not rescue the recipe (λ = 3: 0.865; λ = 27: 0.841). Reinforcement learning also lost at 4B under a cleaner same-start control: a3grpo 0.789, −0.135 (−0.199 to −0.043) below its supervised sibling. Preference optimisation lost less, but still lost: a3dpo 0.881, −0.043 (−0.095 to −0.014) below supervised.

#### Chain-of-thought never paid for itself

The explicit reasoning pass reduced pooled AUROC at 1.5B (−0.072, −0.116 to −0.048), 3B (−0.080, −0.162 to −0.037) and 4B (−0.041, −0.085 to −0.008), and was indistinguishable at 8B (−0.012, −0.043 to +0.009) — a penalty that shrinks with scale but never becomes a gain.

#### Routine-label training tied zero-shot; the mismatch control behaved as designed

The arm fine-tuned on the contaminated routine-label corpus (a0, 0.899) is indistinguishable from the untrained zero-shot baseline (a6z-1.5B, 0.890; Δ +0.009, −0.014 to +0.024): the contaminated supervision bought nothing. The proxy-task control (a1, 0.792) sits far below its own zero-shot base (−0.098, −0.180 to −0.049), confirming that fine-tuning on the wrong label space is worse than not fine-tuning at all.

### 5.3 The frontier model and its two students

The frontier model scores 0.932 pooled (0.886–0.960) — numerically above the regression, but not separable from it (+0.007, −0.005 to +0.026). Its regression student (a5*) scores 0.934 (+0.009 vs. a5_lr, −0.004 to +0.027, n.s.).

Its 4B student is the best arm in the study: **0.940 pooled (0.898–0.963)** — above the incumbent regression (+0.014, +0.004 to +0.031), above the physician-label 4B trained on the *true* reference labels (+0.016, +0.0004 to +0.032), and above its own teacher (+0.008, +0.001 to +0.017). It is also the best-calibrated language-model arm (ECE 0.045, slope 0.86, saturation 16%, against 0.102 / 0.40 / 68% for the physician-label 4B), and the only arm whose fixed-threshold ΔNB100 point estimate (+3.13) exceeds the teacher’s (+2.94). The decision-value contrasts, with intervals and multiplicity correction, are reported in the companion paper [22], where none of the deployable-arm contrasts excludes zero. Two mechanisms are consistent with a student beating its teacher and cannot be separated here: averaging over teacher verbalisation noise (Section 4.3), and the specialist reference standard itself being a noisy routine record.

The label budget saturates early: pooled AUROC is already at the teacher’s level with 50 teacher labels (0.937), reaches 0.940 by 200, and is 0.940 at the full 943 (three-seed spread at budget 472: 0.001 AUROC, 0.19 ΔNB100). The supervision bill for the deployed arm — 943 calls at about 12 s each — is roughly three GPU-free hours of teacher time plus seven minutes of student training.

### 5.4 Calibration separates arms that discrimination cannot

Zero-shot arms discriminate well but are miscalibrated *en bloc*: a6z-4B reaches AUROC 0.929 with ECE 0.309, and the 1.5B, 3B and 8B zero-shot arms flag 100% of the panel at the deployed threshold, collapsing to the trivial refer-everyone policy, while the 4B flags 73%. Fine-tuned arms move probability mass to the extremes: the DPO arm saturates 99% of its outputs, the a2 family 95–98%. The regression, the frontier model, and the distilled 4B are the only well-calibrated arms (ECE 0.039 / 0.041 / 0.045) — and they are exactly the arms whose fixed-threshold decision value is competitive. Training regime, not architecture, is what moved calibration.

### 5.5 Held-out institution

On the held-out site (*n* = 58, 52 cases, any-operator label), point estimates reorder and intervals are wide: zero-shot 3B 0.925, zero-shot 4B 0.894, zero-shot 8B 0.875, frozen regression 0.869, frontier 0.851, distilled 4B 0.843, zero-shot 1.5B 0.808. No shortlisted arm is separable from any other here; the check is reported for transparency, not for selection.

## 6 Discussion

### Audit before you train

The single largest fact about this programme’s routine label — that 40.5% of it was written by accounts that never record a positive — was detectable from two columns of provenance metadata (operator identifier, count, positivity) before any model was trained. The competing-mechanism test matters: had the timestamp hypothesis survived, the remedy (drop bulk-written rows) would have differed from the correct one (drop flagged accounts’ rows, distrust the time column). We suggest operator-level positivity audits as a standard pre-training step for any routine outcome column.

### A strong simple baseline, honestly evaluated, is hard to beat

Every fine-tuned local arm had access to exactly the information the regression uses. None beat it. The pre-registered two-stage recipe — pre-train on the abundant proxy, align on the scarce specialist label — failed in the direction opposite to its motivation. This is consistent with a ceiling set by the instruments and the reference standard rather than by model capacity; the 4B replication (same-start SFT/DPO/GRPO) shows that the RL losses are not a small-model artefact.

### If the frontier model is undeployable, distil it — and count the queries

The frontier model adds nothing as a *predictor* over the regression here, and it cannot be deployed on-site. As a *labelling instrument*, its verdicts, distilled into a 4B that runs on a consumer GPU, produced the best-discriminating and best-calibrated arm in the study — without a single physician label in training, at a supervision cost under one thousand queries. The label-budget curve, the teacher-noise measurement, and the overlap sensitivity are reported so that the recipe can be costed elsewhere.

### What this paper does not claim

Nothing here speaks to reinforcement learning or chain-of-thought on long-horizon reasoning tasks; the GRPO arm’s estimator reduces to vanilla policy gradient with an exact baseline in this two-action setting, and the DPO arm reduces to a logistic margin loss. The distilled arm’s advantage over its teacher is measured against a routine specialist standard, not against adjudicated truth. Pooled numbers are case-mix specific, and the companion paper [22] shows that the evaluation-design terms (outcome definition, clustering) are larger than most model contrasts reported here.

## 7 Limitations

The audit is a lower bound: it detects accounts that never record positives, not subtler default behaviour. The reference standard is a routine physician record — single reader, no adjudication — and its date column is unusable, so temporal precedence is asserted by construction (models postdate diagnoses) rather than verified from timestamps. The development panel is case-mix selected (40.3% impairment); no population performance is claimed. The 1.5B/3B arms are Qwen2.5 while the 4B/8B arms are Qwen3, so cross-size comparisons confound scale with model generation; only within-size contrasts are cleanly interpretable. The held-out site is one case-enriched institution of the same programme. The teacher was not re-queried for self-consistency after a renderer fix, so the stored measurement predates it. Equivalence between the distilled arm and its teacher was not formally tested; statements of “no detected deficit” are interval-based. The effective sample size (~158) bounds the precision of every contrast, and several null results are indeterminate rather than established equalities.

## 8 Conclusion

In one deployed cognitive-screening programme, forty percent of the routine outcome label was default-filled by identifiable operator accounts; training on it bought nothing, and the pre-registered proxy-then-RL remedy was worse than doing nothing. Under a specialist reference standard and leakage-free clustered evaluation, no fine-tuning, preference-optimisation, or reinforcement-learning recipe on a few hundred specialist cases beat a 21-variable logistic regression. What did beat it on pooled discrimination — with the best calibration in the study and decision value at least matching every alternative — was distilling fewer than one thousand frontier-model verdicts on the programme’s own unlabelled records into a locally deployable 4B model. The audit, the arm matrix, and the distillation recipe are released as frozen, script-regenerable artefacts, shared with a companion paper that audits the evaluation itself.

## Data Availability

All data produced in the present study are available upon reasonable request to the authors. The de-identified analysis dataset supporting the findings, together with all analysis code and derived artifacts, is available from the corresponding author on request. The programme's identifiable source records are held by the data custodian (Beijing Medical Award Foundation) and cannot be redistributed.

## Declarations

### Ethics

Individuals gave informed consent electronically through the programme’s information system when receiving services at the participating institutions. The study — a retrospective statistical analysis of existing, de-identified programme records, with no clinical intervention — was approved by the Ethics Committee of the Medical College of Qingdao University (approval no. QDU-HEC-2025431, 22 May 2025), and conforms to the Declaration of Helsinki [21].

### Data availability

The de-identified dataset supporting the findings is available from the corresponding author on request, as are all analysis code and derived artefacts. The programme’s identifiable source records are held by the data custodian (Beijing Medical Award Foundation) and cannot be redistributed.

## Acknowledgements

The authors thank Tao Li, Haifeng Zhang, Huizi Li, and Yuhan Xie (Dementia Care and Research Center, Peking University Sixth Hospital, Peking University Institute of Mental Health, Beijing) for their support in study coordination and data curation; Mengmeng Xia (Beijing Jiahua Social Work Service Center, Beijing) for programme administration; and all institutions participating in the programme.

## Generative AI declaration

During the preparation of this work the authors used Claude (Anthropic) extensively and under author direction for writing and revising the analysis and training scripts, for building verification tooling that cross-checks manuscript numbers against the frozen result files, and for drafting and revising manuscript text. All analyses, numbers, and claims were reviewed and verified by the authors against the frozen result files, and the authors take full responsibility for the content. The language models evaluated in this study are objects of study, not writing tools, and are reported in the Methods.

